# Factors affecting patients on antiretroviral therapy lost to follow up in Asunafo South District of Ahafo Region, Ghana: Cross-sectional Study

**DOI:** 10.1101/2024.01.17.24301449

**Authors:** Robert Kogi, Theresa Krah, Emmanuel Asampong, Edward Mberu Kamau

## Abstract

**Background:** Despite the increased and effective program coverage for antiretroviral therapy (ART), a considerable proportion of individuals receiving ART discontinue medication at different stages of their treatment pathway. In sub-Saharan Africa, approximately half of individuals who test positive for HIV are lost to follow-up. This study was set out to answer the following question “What are the factors that affect patients on ART loss to follow-up in Asunafo South District of Ghana?”

**Methodology:** Cross-sectional study design with systematic random sampling was employed to select HIV patients on ART and those lost to follow-up. Stata 17.0 was used to analyze the data. A cox-proportional hazard regression was fitted in order to determine the predictor variables. Variables for the multivariable cox-proportional hazard regression model were chosen by entering the outcome variable (loss to follow-up) and explanatory variables into the model. Lastly, the association between the explanatory and outcome factors was determined using the adjusted hazard ratios and their associated 95 percent confidence interval considered.

**Findings:** Patients who began antiretroviral therapy at age 41 years or older had a significantly lower chance of being lost to follow-up than those who began ART at age 35 or less. Furthermore, patients who started ART with a primary education had 1.68-fold increased risk of lost to follow-up compared to patients with no education. In addition, patients in rural locations had a 2.65-fold higher likelihood of being lost to follow-up than patients in urban areas. The main reasons for missing ART appointments among patients included walking long distance to clinic, cost of transportation, fear of scolding from clinic staff, stigma, and erratic supply of antiretrovirals. In conclusion, to reduce HIV patients lost to follow-up, all clinicians and stakeholders should take into account the risk factors that have been identified when providing ART services and counselling.

**Key Messages:** The existing knowledge on the topic is that despite increased and effective program coverage for antiretroviral therapy, a substantial proportion of individuals discontinue medication during various stages of treatment, particularly in sub-Saharan Africa, where approximately half of HIV-positive individuals are lost to follow-up or do not undergo eligibility assessments for therapy after testing positive.

As a result of this study, we now know that initiating antiretroviral therapy at age 41 or older, having no formal employment, residing in urban areas, and possessing a higher education level are associated with a significantly lower risk of lost to follow-up among HIV patients, and addressing factors such as long-distance travel, transportation costs, fear of stigma, and antiretroviral supply issues are crucial in reducing patient attrition from ART.

Finally, this study emphasized the importance of considering these factors in providing comprehensive ART services and counseling to reduce HIV patient disengagement from ART.

## Introduction

The World Health Organization (WHO) has observed that HIV remains a global public health concern [1]. Despite the fact that HIV infection cannot be cured, persons living with the virus can now live long and healthy due to improved access to HIV prevention, diagnosis, treatment, and care [2]. Regardless of clinical status or CD4 cell count, all individuals living with HIV, including children, adolescents, adults, and pregnant and lactating women get lifelong antiretroviral therapy (ART) [3].

There were 342,307 estimated people living with HIV (PLHIV) in Ghana as of 2019, with a total estimated 20,068 new infections [4]. Moreover, the estimated ART coverage among adults (15+ years) was 46.55% [4].

The World Health Organization’s “treat all” policy, which was previously utilized as a cutoff point to begin treatment, was accepted by the Ghanaian government [5]. As a result, efforts to maintain the long-term effects of ART and lower the risk of new HIV infections are receiving more attention in Ghana [6]. However, in sub-Saharan Africa patients typically have low retention rates [7, leading to LTFU.

The antiretroviral coverage in Ghana is estimated to be at 45% among PLHIV [8]. The immunological benefits of antiretroviral medication are negatively impacted by loss to follow-up, which also increases hospitalizations, morbidity, and death from AIDS among HIV patients [9].

Additionally, it was discovered that patients in Lagos, Nigeria, provided false contact details out of a refusal to accept HIV-positive results and to enrol in treatment, as well as a fear of stigma associated with the virus [10]. In Ghana, not much research has been conducted to investigate the precise causes of HIV patient loss from antiretroviral therapy (ART), particularly in the Asunafo South District of the Ahafo Region. In the Asunafo South District of the Ahafo Region, report on the District Health Management System (DHIMS2) for 2020 and 2021 showed that out of 3,090 and 4,017 HIV patients who were on ART, some 1,593 and 1,564 patients have respectively stopped treatment due to loss to follow up [11]. Knowing why people stop taking antiretroviral medication (ART) is critical because maintaining patients on ART and ensuring adherence to therapy are critical components in achieving successful long-term outcomes. Therefore, in order to help patients on ART stay on treatment, we carried out this study to answer the question “What are the factors that affect patients on ART loss to follow-up in Asunafo South District of Ghana?”.

### Conceptual Framework

This study sought to construct an overarching conceptual model to guide in the understanding of HIV patient lost to follow-up in the Asunafo South District. The Andersen and Newman Behavioural Model (ANBM) for health service utilization was used. The Andersen Newman Framework [12] posits that an individual’s access to and use of healthcare is a function of three main factors: 1) Predisposing factors (socio-cultural characteristics of individuals that exist prior to their illness); 2) Enabling Resources (the logistical aspects of obtaining care, which can include personal, family and community resources); and 3) Need factors (the most immediate cause of healthcare use from problems that generate the need for care).

This framework was used because, it has both predictive and explanatory capabilities and provides insight on how to maintain, or in some cases improve, the health status of the populations under study. Additionally, the grouping of factors under study as predisposing characteristics, enabling resources and need factors can guide the development of appropriate interventions relevant to the study problem. While many predisposing characteristics such as age may be difficult to alter, to change utilization, certain enabling resources such as type and location of care and need factors, particularly those relating to perceived needs for care, may be easier to address [12]. Moreover, the framework allows for the consideration of broad environmental factors. Although we may not be able to measure the direct role of ART policies and tracing guidelines in this research, the framework was intended to help to identify mutable factors under study that could be affected through key policy and program changes to reduce losses to follow-up in the district. In addition, this framework has also been used as a behavioural model for vulnerable populations, by specifically placing greater emphasis on environmental enabling resources and need factors [13].

Primarily, this study was interested in identifying how various factors influence health behaviours with regard to LTFU. Though they are difficult to measure directly, we recognize the importance of more distal acting environmental factors, and their potential impact on health behaviours and outcomes. Hence, this conceptual model based on the framework was pertinent to the research objectives of this study.

## Materials and Method

### Study Design

A facility-based cross-sectional study was used in this study. The data collection was conducted using the facilities’ records and tracing respondents for interview.

### Study Location/Area

The study was conducted in the Asunafo South District in Ahafo Region of Ghana. According to the 2020 population and housing census, the district has a total population of 93,619 with a total number of children 0-11 months and women in reproductive age being 3,668 and 22,006 respectively [14].

In terms of health service delivery, Asunafo South District is divided into four (4) sub-districts; Kukuom, Sankore, Kwapong and Abuom with twenty health facilities of which twelve (12) are functional CHPS distributed throughout the entire sub-districts. The district has two HIV clinics rendering ART services (Asunafo South District Hospital and Sankore Health Centre). Approximately 4,017 clients were actively receiving ART by the end of 2021, and the HIV clinics are open five days a week. Patients from all backgrounds and communities inside and outside the district are served by the clinics.

Figure 1 illustrated the district map indicating the ART sites.

**Figure 1:**
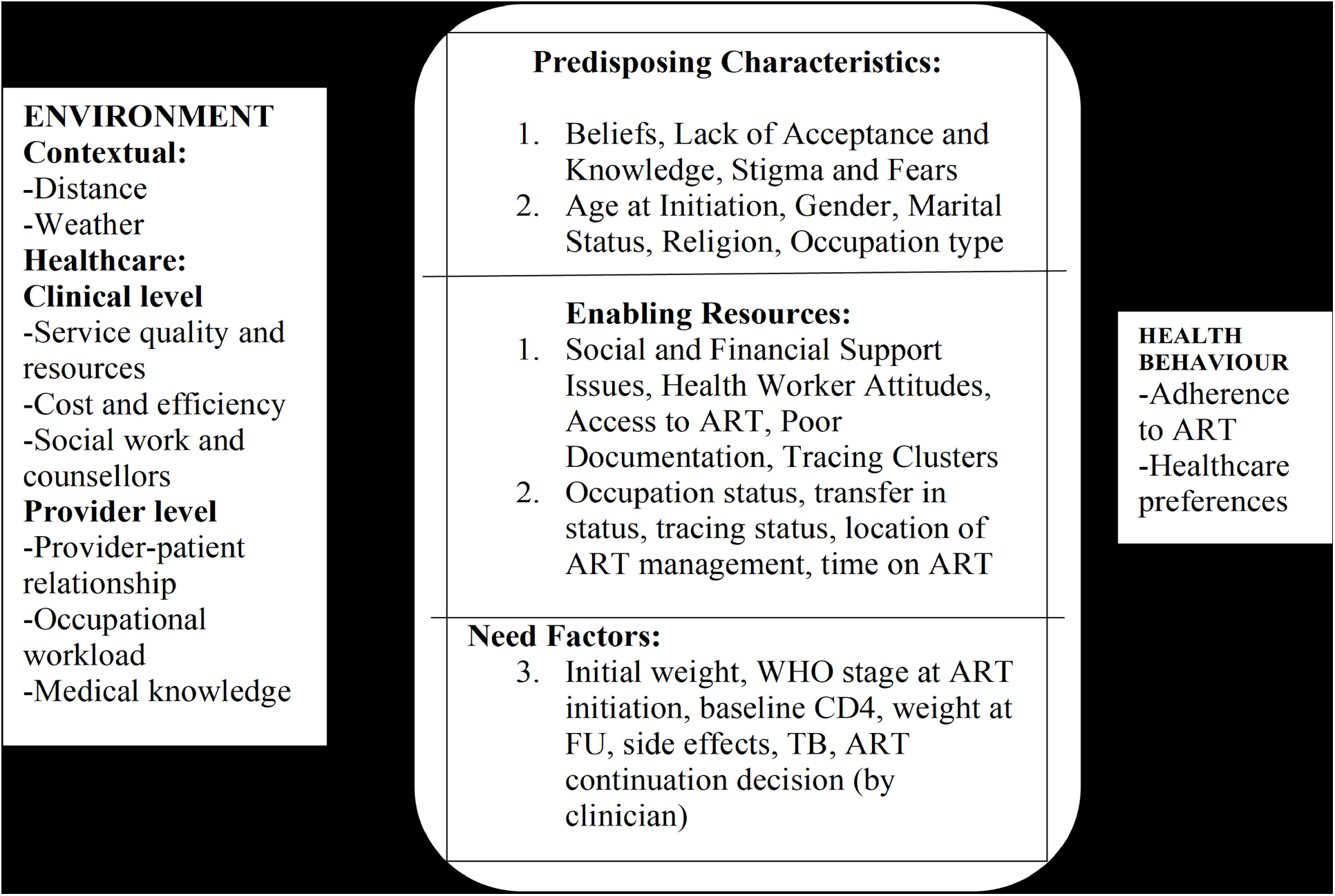
Conceptual framework of potential factors associated with becoming LTFU from ART.

**Figure 2:**
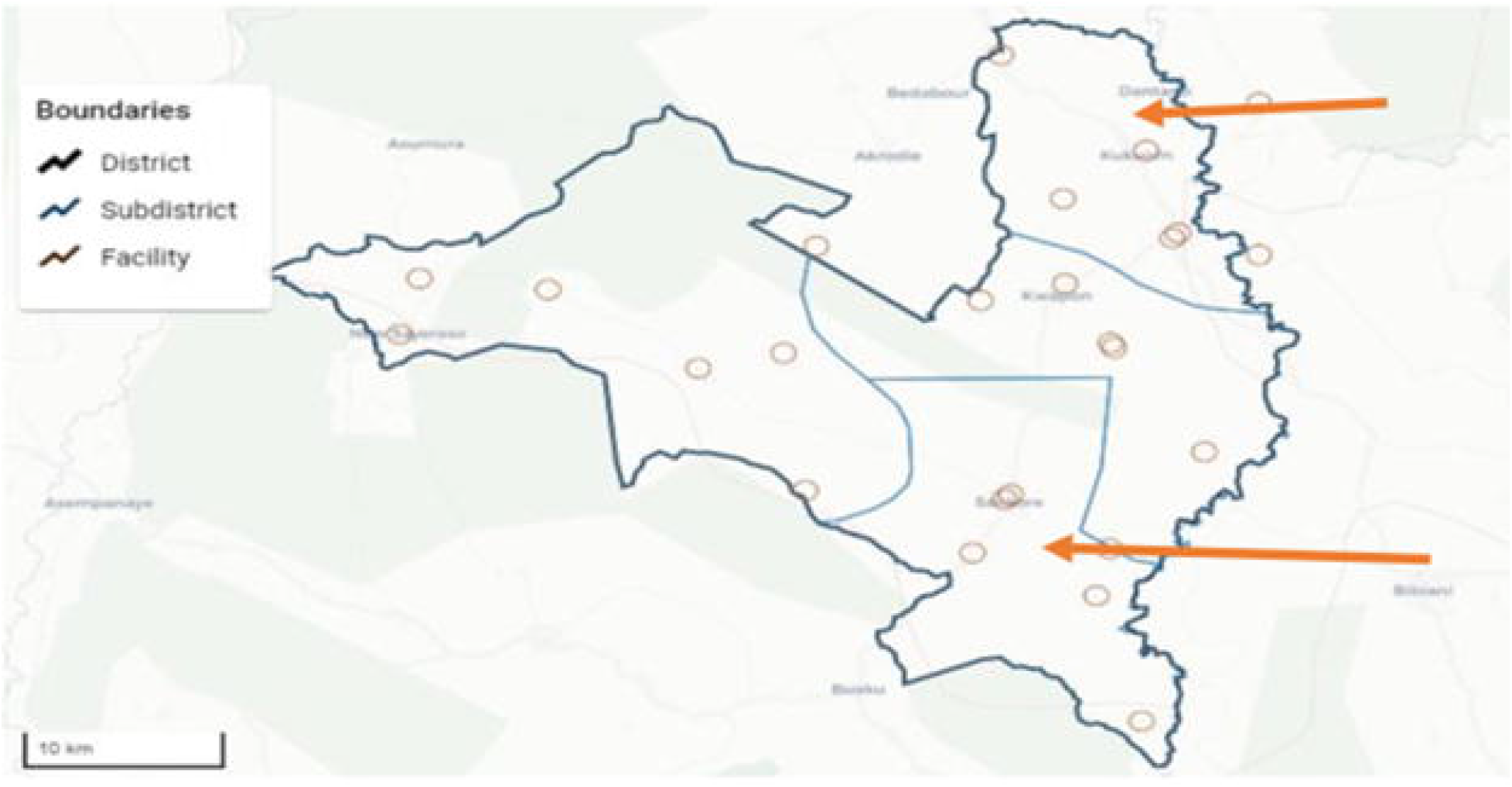
Map of Asunafo South District.

### Variables

The outcome variable in this study was LTFU from ART services, whereas the sociodemographic variables included in the analysis were sex, age, education level, marital status, occupation status, and place of residence, the quantity of tablets taken daily, the level of adherence, and adverse drug reactions were all regarded as independent variables.

### Study Population

The study population included individuals who were on ART as at the end of 2021 or were no longer in care and have been lost to follow-up. Complete location information (10-digit phone number) in the HIV testing and counselling registration was the inclusion requirement for tracing. A minimum of 18 years of age and prior experience with ART as a patient were prerequisites for inclusion.

### Sample Size

The Cochran formula was used to determine the study’s sample size [15]. The standard normal (Z) value corresponding to the 95 percent confidence interval, a precision level of +/− 5 percent, and an expected retention level of 72 percent were the parameters employed in this sample size estimation [14].

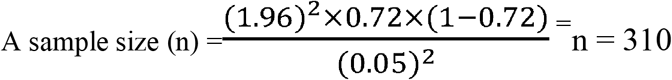

In order to account for clustering in LTFU at the health facility level, the sample size calculated by this method (N = 310) is inflated by a design effect of two, resulting in a total sample size of 620 [16].

### Sampling method

Systematic random sampling was used to select HIV patients on ART and those lost to follow up. The first patient in this systematic sampling was chosen at random, and subsequent patients were then drawn from the clinics database (registers) in accordance with a sample interval defined as 5,581/620 (i.e., every ninth patient in each ART clinic’s register) until the targeted sample size was reached. The sample size was achieved by using the clinics register as a sampling frame.

### Data Collection Techniques/Methods & Tools

Five (5) data collection assistants with expertise in gathering data for public health research were contracted and given training on how to collect the data. First, eligible HIV patients on ART and those on ART who stopped responding to follow-up calls were contacted to explain the objectives of the study and set up appointments for in-person structured interviews or phone interviews for those who could not be reached but were willing to take part in the study. It took at least three times to make phone calls on several days and at various times of the day to conclude that the tracing was unsuccessful.

The data was gathered using a questionnaire that was created on the KoboCollect platform. This reduced the amount of time that respondents had direct contact with the questionnaires and prevented the possible spread of COVID-19. A standardized and pre-tested questionnaire was used to gather the data; it was created in English and, for those who could not understand it, was translated into Twi by data collection assistants.

Data collection and patient tracing were carried out in August and September, 2022. Each interview lasted from thirty to sixty minutes.

### Quality Control

The data collection process was carefully undertaken to ensure the quality of the data, and we ensured that data collectors who were selected had prior data collection experience. Prior to data collection, a pre-test was administered to ten (10) ART patients in the Asunafo North Municipal on populations that were similar. Throughout the whole data collection period, the investigators conducted intensive supervision on a daily basis. Before the data collection period began, the investigators examined the data collection tool to ensure its dependability. We also cross-checked the data at the end of each day to ensure it was accurate, consistent, and complete. As a result, a corrective discussion was held with each data collector. During discussion periods, different views were considered in an effort to reduce errors and corrective measures implement. After that, the data were coded, entered into the computer, and examined for consistency and completeness.

### Data Analysis

Excel was used to extract and verify the completeness of the data from the KoboCollect data collection application. After that, it was exported to Stata 17.0 for processing, cleaning, editing, and analysis. Each study respondent’s results were classified as either censored or event-related (LTFU). Additionally, a cox-proportional hazard regression model was applied in order to determine the predictor variables.

Frequency distributions or percentages were used to describe the categorical data that was taken from HIV-positive clients. In terms of bivariate analysis, variables for the multivariable cox-proportional hazard regression model were chosen by entering the outcome variable and explanatory variables into the cox-proportional hazard regression model. Thus, the multivariable cox-proportion regression model was fitted to the variables in the bivariable analysis with p-values ≤0.25. Lastly, the existence of a meaningful association between the explanatory and outcome factors was determined using adjusted hazard ratio and its associated 95% confidence interval (CI). Significant variables were those with p-values less than 0.05.

### Statistical Methods

The statistical methods which were used in this research included descriptive statistics, cox-proportional hazard regression model, univariate, bivariate, and multivariate analyses.

### Ethical Consideration

We sought ethical approval from the Ghana Health Service Ethics Review Committee (GHS-ERC), with approval number: **GHS/ERC/021/06/22**. Permission was also obtained from the Ahafo Regional Health Directorate and Asunafo South District Health Directorate before we conducted the study. Informed consent was sought from study respondents in a written form, where the consent form was given to each participant to read or get someone, they trusted to read for them before they participated. Two copies of the form (one for each participant and one for the research team) were given to each participant the endorse. The participants were assured of their confidentiality and anonymity of the information they provided.

## Results and Discussion

### Results

#### Description of the study respondents

The results in Table 1 showed that majority of the respondents in this study were at least 46 years of age, 197 (32.83%), followed by respondents who were 31-35 years old, 125 (20.83%). The mean of respondents in this was approximately 40.71, with a standard deviation (S.D) ±13.55. the minimum and maximum ages of respondents were respectively 21 years and 77 years. Majority of the respondents were females, 344 (57.33%) and a greater proportion of the respondents attained Junior and or Senior High school education, 372 (62.00%). A greater number of the respondents were Akans, 429 (71.50%). In addition, majority of the respondents were Christians, 526 (87.67%). Respondents who were married at the time of the conducting this study formed the greater majority of 231 (38.50%), followed by those who were single, 164 (27.33%). Also, a majority of the respondents who participated this study were farmers, 279 (46.50%), while most of them indicated they were urban dwellers, 459 (76.50%).

**Table 1: Socio-demographic Characteristics of Respondents at the Asunafo South District**

#### Use of antiretroviral among study respondents

From Table 2, it is shown that majority of the respondents were on ARVs, 564 (94.00%) at the time of conducting this survey. Among those respondents who were on ARVs, a greater proportion of them were taking first (1^st^) dose (First line), 564 (94.00%). It is further shown that majority of the respondents who were on the ARVs indicated they regularly took their drugs, 534 (94.68%). For the respondents who were on the ARV, a greater number of them said they took from the clinic, 549 (97.34%). Finally, though a majority of the respondents indicated they never missed their ARVs, 387 (68.62%), some 127 (22.52%) and 28 (4.96%) said the last time they missed their ARVs were respectively 1-4 weeks and more than 3 months ago prior to the conduct of this survey.

**Table 2: Use of antiretroviral among ART patients**

#### Reasons for discontinuing ARV medication

The results in Table 3 showed that in terms of access to care, the major reasons why some respondents may discontinue care were; walking long distance to clinic, 352 (58.67%) and high cost of transportation, 286 (47.67%). In terms of clinic quality, majority of the respondents discounted the assertion that staff were not nice, 537 (89.50%), were afraid to be scolded by clinic staff, 586 (97.67%), and that attending clinic did not risk disclosure of their status to community, 570 (95.00%). It is however shown in the results that many of the respondents noted that feeling healthy, 332 (55.33%) was one of their personal reasons why they may not continue with the intake of ARVs. Though majority of the respondents said they needed the ARVs, 558 (93.00%), some 42 (7.00%) indicated that they didn’t need it. Also, most of the respondents refuted the assertion that they were not permitted by religion or faith to continue with the intake of their ARVs, 571 (95.17%). However, some proportion of 29 (4.83%) confirmed that were not permitted by religion or faith to continue taking the ARVs.

**Table 3: Reasons for discontinuation of care among ARV patients**

#### Predictors of patients Lost to Follow Up

Table 4 depicts the unadjusted, full adjusted and final adjusted hazard rates of factors associated with LTFU. Patients with primary education were approximately 4.0 times more likely to be lost to follow-up at the full adjusted hazard model level (aHR^a^ = 4.01, 95%CI; 1.15-6.40). Male patients, on the other hand, were found to be significantly 94% less likely to be lost to follow-up (aHR^a^ = 0.06, 95%CI; 0.01-0.33), and being a Fante client was found to be 93% less likely to be lost to follow-up (aHR^a^ = 0.07, 95%CI; 0.01-0.85). Additionally, compared to respondents who were single, married patients had a significant 73% lower chance of being lost to follow-up (aHR^a^ = 0.27, 95% CI: 0.08-0.86). Furthermore, respondents who did not work in the government sector had an 81% lower chance of being lost to follow-up (aHR^a^ = 0.19, 95% CI; 0.07-0.52). Respondents who resided in rural areas had a much higher chance of being lost to follow-up compared to patients who lived in urban settings (aHR^a^ = 6.82, 95%CI; 2.49-18.72).

In the final cox’s proportional hazard model, the risk of LTFU was shown to be considerably lower among respondents who started antiretroviral therapy at age 41 or above compared to those who started ART at age 35 or below (aHR^b^ = 0.34, 95% CI, 0.13-0.84). Compared to respondents without any education, those who began antiretroviral therapy and had only completed primary school had a 1.68-fold higher chance of dropping out of the study (aHR^b^ =1.68; 95%CI; 0.83-3.43). Comparing respondents who were unemployed to those who were employed, there was a 69% reduction in LTFU (aHR^b^ = 0.31, 95%CI; 0.14-0.69). Patients who lived in rural areas were 2.65 times more likely to get lost to follow-up care than patients who lived in urban areas (aHR^b^ = 2.65, 95% CI, 1.29-5.44).

**Table 4: Hazard ratios of patients lost to follow-up being unadjusted and adjusted among 600 respondents**

## Discussion

The purpose of this study was to identify the factors that affected ART patients’ failure to remain on ART at public health institutions in Asunafo South District. Of the 620 targeted sample size for this study, 600 patients representing about 96.8% who participated had their data fully completed and qualified for consideration in the analysis. More than half (57.33%) of the respondents in this study were females. This result is consistent with the national trend of women having a comparatively greater prevalence of HIV and AIDS than men [17]. This finding may have some bearing on the mandated elimination of mother-to-child transmission of HIV (e-MTCT), in which women are tested as part of standard antenatal care (ANC) services in Ghana [18].

The finding from the study suggests that while a majority of the respondents were on antiretroviral therapy (ARV) for HIV treatment, a small percentage of them (6.0%) were not taking their medications regularly. This indicates the presence of a gap in adherence to ARV treatment among some HIV patients. The result is in line with what was reported in Southern Ethiopia which found that 9.1% of HIV patients did not continue receiving treatment after starting it [9]. However, it differs from a study by [18] that found more than half (55.6%) of their study respondents were not on antiretroviral therapy.

Among the respondents who were on ARVs in the present study, a majority of them were taking the first line (94.00%) and were regularly taking their drugs (94.68%). Moreover, most of the respondents preferred to receive their ARVs from the clinic (97.34%), and this preference is consistent with a finding from Ghana that the majority of PLHIV preferred to acquire ART drugs from medical professionals, and the hospital was the most popular location for these prescriptions [18]. The preference of patients for receiving ARVs from the clinic and health personnel in this study indicates the importance of ensuring adequate access to these resources for HIV patients.

The findings of this study further highlighted some important factors that contributed to discontinuation of care and non-adherence to ARVs among HIV patients. These include barriers to access to care such as long distance to clinics, high cost of transportation, and long waiting times. We found that some respondents were concerned about the quality of care, including concern of being badly treated by health workers (10.50%), others were also afraid of scolding from clinic staff (2.33%), while some patients said attending the clinic risked disclosure of their status to the community (5.00%). Other research expressed a similar concern, stating that maintaining a good rapport with the clinic staff was essential to keeping patients there [20,21]. This current study’s findings also supported what was reported in southern Mozambique, that the most frequently reported barriers to patients lost to follow-up were the fear of bad treatment from health personnel [22].

Furthermore, the results of this study showed that a little more than half of the respondents (55.33%) stated that they were not ready to continue taking ARVs since they were feeling well, and almost 7.0% of them felt they did not need the ARVs at all, because they were feeling healthy. This finding corroborated with what was found in southern Mozambique, that one of the most frequently reported barriers to patients lost to follow-up was the perception of being in good health [22].

Also, though most of the respondents refuted the assertion that they were not permitted by religion or faith to continue with the intake of their ARVs in this study, it was found that a proportion of about 4.8% confirmed that they were not permitted by their religion or faith to continue taking their ARVs. Other respondents (approximately 1.2%) have ever sought HIV treatment from alternate sources. A similar finding was reported that patients believed that God can ‘cure HIV’ and sought their care from different places instead of the hospital [23]. This could be affecting clients’ readiness and adherence to the ARVs medications. A similar finding was reported in Kumasi-Ghana, that the study respondents believed alternative or herbal remedies could cure their HIV infections [24].

### Predictors of clients Lost to Follow-Up

Respondents who started ART at age 41 and above had a much lower risk of long-term follow-up than patients who started ART at age 35 or below. This difference in risk was statistically significant. This result suggested that, in comparison to older age groups, younger patients were more likely to be lost to follow-up from ART. One possible reason could be that younger patients are more likely than older patients to be mobile and to fear stigma associated with HIV [24]. This may also be related to patients’ immaturity in their ability to think critically and the unique difficulties that come with being younger than older patients. This was in agreement with what was reported that younger age could results in lost to follow-up and that age less than 35 years was a predictor of LTFU from ART by [25,26]. Also, patients who started ART with a primary education level had a 1.68-fold increased risk of LTFU compared to those without any education. This agreed with the findings reported in another study that atherosclerosis in pre-ART care may be exacerbated by decreased educational attainment [27]. Furthermore, a little above two-third of patients who did not work for the government had a significantly lower likelihood of becoming lost to follow-up than patients who were unemployed. This implied that patients who were employed in one way or the other may be a bit financially stable and may perceived seeking care at different places and sources as compared to the unemployed who may lack money, and had no option than to ensure regular follow-up for their ART. This supported what was reported in North-western Ethiopia that lack of employment was a strong predictor of not receiving ART [28]. In this present study, we also discovered that patients were 2.65 times more likely to be LTFU in rural areas than they were in urban settings. This may be the result of issues with accessibility or distance from treatment facilities, transportation expenses, a lack of knowledge about the advantages and risks of treatment adherence, and societal stigma associated with HIV and AIDS among patients from rural areas.

## Conclusion

This current study has added to the existing literature ideas on factors affecting HIV treatment. First, compared to patients who started ART at a younger age, this our study revealed that patients who started ART at an older age (41 years or above) had a considerably decreased risk of LTFU. This provides a crucial insight, emphasizing the necessity of focusing treatments on younger patients in order to increase adherence to antiretroviral therapy. Second, the finding that patients with only primary education were more likely to experience lost to follow-up than patients with no education points to the need of health education programs to increase low-education patients’ adherence to antiretroviral therapy.

Furthermore, it was demonstrated that patients who did not work for the government had a lower likelihood of lost to follow-up than the unemployed. A noteworthy finding that emphasizes the need to address the particular difficulties rural patients encounter in getting and sticking to ART is that they were most likely to terminate their ART treatment than patients who lived in urban areas. Finally, this study has identified various reasons for missing ART appointments, such as the cost of transportation, stigma, and perceived side effects of the drugs, provided valuable insights into the factors that influenced patients lost to follow-up from ART.

Therefore, all physicians should take into account the risk factors that have been identified in this study when providing ART services and counselling.

### Policy recommendations

1. Ghana AIDS Commission in collaboration with Ghana Health Service should develop strategies to include interventions to address the concerns of patients about the quality of care, fear of stigmatization, and other barriers to access care.
2. Ghana AIDS Commission in collaboration Ghana Health Service should train health personnel to provide quality care and support, and providing psychosocial support services to address the fears and concerns of patients.
3. Ghana AIDS Commission, Ghana Health Service, and other stakeholders should develop targeted interventions to address the needs of younger patients and patients with lower levels of education who are at a higher risk of LTFU. Strategies could include improving education and awareness about HIV and ART, providing support for transportation and other related costs, and improving access to care for patients who are unemployed or have low incomes.

## Limitations of the study

Our study was constrained by the fact that, in order to account for potential bias, we did not compare patients’ missing information to those who did not, on important characteristics associated with LTFU. Furthermore, a few factors that may have been overlooked, such as having a caregiver, having a viral load, having a mental disorder, and body mass index, could also be predictive factors for lost to follow-up. This study may have underestimated or overestimated the incidence rate of loss to follow-up because it did not take into account moved out patients and inadequate records.

## Acknowledgement

We are grateful to Ahafo Regional Health Directorate and Asunafo South District Health Directorate for granting us the permission to conduct this study. We also thank the facilities’ In-charges and ART nurses for their assistants in getting the needed study respondents. To our research data collection assistants; our research team, and especially all communities and participants involved, we say thank you.

## Contributorship statement

RK conceptualized the study and planned its conduct with TK and EA. All the authors wrote the introduction, reviewed the literature and wrote the methodology. RK sought ethical clearance from the Ghana Health Service Research Ethics Committee. RK and TK collected the data, and RK carried out the analysis. The reporting of the work was done by all the authors. The final version of the manuscript was written, reviewed and accepted by all the authors.

The corresponding author attests that all listed authors meet authorship criteria and that no others meeting the criteria have been omitted.

## License

The Corresponding Author has the right to grant on behalf of all authors and does grant on behalf of all authors, an exclusive licence (or non-exclusive for government employees) on a worldwide basis to the BMJ Publishing Group Ltd (“BMJ”), and its Licensees to permit this article (if accepted) to be published in The BMJ’s editions and any other BMJ products and to exploit all subsidiary rights, as set out in our licence.

## Data Availability Statement

The data for this study is available upon official request from the corresponding author through

